# Analysis of Time-to-Death Survival Function in a Nationally Representative Random Sample of HIV-seropositive Treatment-experienced Adult Patients from Malawi – A Historical Cohort Sample of 2004-2015 HIV Data

**DOI:** 10.1101/2024.03.13.24304062

**Authors:** Hemson Hendrix Salema

**Author notes:** Corresponding author:, (alt.).

## Abstract

How rapid HIV infection progresses to AIDS and to death is affected by different factors. This study explores survival times and associated survival factors from treatment initiation to death or censoring in antiretroviral therapy-experienced HIV-seropositive adults in Malawi from 2004-2015.

A multicentre non-concurrent, retrospective cohort study was undertaken from eight ART Centres where patients’ medical records (PMRs) of HIV-positive adult patients aged 15+ years were reviewed. A life table, the Kaplan-Meier log-rank, and Cox Proportion Hazard regression were used to calculate survival time-to-death and its correlates, respectively. Hazard ratio with 95%CI and p<0.05 were used to declare statistical significance.

Data for (n=9,953) patients were abstracted from PMRs. Patients median age was 40 (IQR: 33-48 years). 60.8% were females, 45.2% were aged 20-39 years, and 78.8% were married. At treatment initiation, 48.1% had advanced HIV disease clinical stage III, 24.5% had WHO stage IV, whereas 27.5% were asymptomatic – of which, 24.9% and 2.6% initiated ART due to low CD4+ count and PMTCT’s Option-B+ eligibility criteria, respectively. Survival function findings revealed that each patient had a single entry into the study. Exit time ranged from 1 to 9,224 days with the mean value of 2,421.9 days, occurring at the rate of 0.00004883 event-failure per-person-day. Time-to-death was observed at the rate of 1.78/100 person-years-at-risk (PYAR). 213 deaths (18.1%) occurred early in year-one post-ART-initiation. Deaths occurred more among persons of 20-39 years (N=470, 39.97%), and of 40-54 years (N=483, 41.07%), and was mostly due to mycobacterial pathogenic conditions (N=106, 37.3%) in particular TB infection (N=103); most of which were PTB cases (N=69, 66.9%). Mortality was high in Southern region (63.1%, N=743) but was least in Northern region (N=313) [*p<*0.0001]. In a multivariate Cox regression predictive model, males gender (aHR=1.42), patients age-groups of 20-39 years (aHR=1.63), 40-54 years (aHR=1.71), and 55+ years (aHR=2.66), Mzuzu Central hospital ART centre (aHR=2.66), Thyolo District hospital ART centre (aHR=3.02), semi-rural areas (aHR=1.30), urban areas (aHR=0.80), being single (aHR=0.86), chronic cough and/or breathlessness (aHR=1.19), chronic diarrhoea or weight loss (aHR=1.43), chronic fever and/or severe headache (aHR=1.30), skin or oral lesion (aHR=1.33), WHO clinical stage III (aHR=17.90), WHO clinical stage IV (aHR=20.09), low baseline CD4 count <250 cells/µL, (aHR=1.17), high baseline VL>1,000 copies/mL (aHR=2.46), Nevirapine-based therapies (aHR=1.14), and HIV duration of 3-5 years (aHR=1.17), 6-10 years (aHR=1.19) and >10 years (aHR=1.16) were all statistically significantly associated with time-to-death.

This study has demonstrated survival factors associated with time-to-death among HIV-positive adults in Malawi. In order to effectively reduce AIDS mortality and win the war against AIDS-related death, the need to critically address and carefully prioritise the identified factors in HIV/AIDS management is great and cannot be overemphasised.

## BACKGROUND

Since discovered, the human immunodeficiency virus and acquired immunodeficiency syndrome (HIV/AIDS) remain a socioeconomical and clinical challenge and highest leading infectious cause of mortality and morbidity worldwide [1, 2]. Though a global challenge, low- and middle-income countries (LMICs) remain disproportionately affected and the sub-Saharan African (SSA) region has been worst hit [2]. HIV/AIDS prevalence has been persistently and constantly higher. Cumulative figures indicate that up to 85.6 million people have been infected with HIV [1, 2]. The recent 2023 HIV report by the World Health Organisation (WHO) and the Joint United Nations programme on HIV/AIDS (UNAIDS) states that globally, the number of people living with HIV (PLHIV) at the end of 2022 was an estimated 39 [33.1–45.7] million – of which, 25.6 million [21.6–30.0 million] were in the African region (SSA) whilst Eastern Mediterranean Region had lowest prevalence of 490,000 [420,000–600,000] [3].

The HIV virus; with its complex pathophysiology that demands complex treatment regimes, remain incurable, though chronically treatable [1, 2]. HIV is naturally highly immunosuppressive and capable of progressively destroying one’s immunity as the disease progresses, eventually leading to imminent death. Since the start of the epidermic an estimated 40.4 [32.9-51.3] million people have died of AIDS-related causes [2]. Of the total global AIDS mortality, most deaths occurred in the African region; in particular, the SSA region with rates up to 292.95 deaths/100,000 people (in Mozambique) [3], than any other regions [1] and, females remained disproportionately affected with rates as high as 70.6% in Congo [3]. Death from HIV by age clearly indicates that 15-49-year-olds have been consistently affected. For example in 2004 when HIV/AIDS mortality was at highest peak; 1.31 million deaths occurred among the 15–49-year-old population compared to 251,698 and 24,537 deaths in adults of 50-69 years and 70+ years old [3]. Recent data shows that 630,000 [480,000–880,000] people died of AIDS in 2022 [2] – the least recorded AIDS mortality in history of HIV/AIDS as opposed to 1.84 million high annual death of 2004 [3].

Death in HIV-seropositive individuals occur as the disease progresses from the acute phase to the advanced HIV disease (AHD) stage; the acquired immunodeficiency syndrome (AIDS), through the (usually protracted) chronic asymptomatic phase. The AHD is marked with progressive increase of viral-RNA load (VL) and a corresponding decrease in body’s immune defence system – manifested by depletion of CD4+ cell count. If untreated or ineffectively treated, the physiological impact on these HIV surrogate markers; thus, CD4 cell count and viral-RNA load, will then lead to physical manifestation of the WHO-classified clinical stages III and IV symptoms; – which are severe stages of HIV infection called AIDS which eventually lead to death. How rapid HIV infection progresses to AIDS varies and can be affected by different host and non-host factors, as are survival times from HIV infection to death, or from treatment initiation to death.

Previous epidemiological evidence has established independent factors such as gender, age, CD4 count, viral load levels, baseline weight & body mass index (BMI), opportunistic infections status, comorbid conditions, ART regimen and adherence levels, ethnicity, poverty, illiteracy, inequality, geographical location, bio-clinical factors, route of HIV infection, diet, pregnancy, stress, cigarette smoking or recreational drugs usage, and healthcare provider experience in managing HIV patients, can both influence the rate of disease progression in HIV patients but also as mortality risk factors [4-10]. However, evidence to that effect from Malawi is lacking. This study aims to explore survival times from ART treatment onset to death or censoring (measured in days) alongside associated survival factors in Malawi using the sub-cohort sample from the large ADROC research study. The study’s outcome variable is survival time in days, the event (or failure) factor is ‘death’, and the response time is the time when patient died.

## METHODS AND MATERIALS

### Study Design, Setting, and Participants

A multicentre non-concurrent, retrospective cohort study was conducted in 2016 between March and May in Malawi from eight ART Centres where patients’ medical records (PMRs) of HIV-positive adult patients aged 15+ years old were reviewed. Health facilities offering ART were randomly selected using multistage probability. HIV positive patients’ medical records called “ART Master cards” were reviewed as sampling frames. Patient’s register and clinical progress notes were consulted to complement for missing or additional information if necessary. Data collection sites included all existing levels of health facility establishments in Malawi: primary, secondary, and tertiary levels. Follow-up time for each patient was calculated from treatment-initiation date to date of death.

### Study population: Inclusion & Exclusion Criteria

Only HIV positive patients aged 15+ years who were once commenced on ART following standardised treatment protocols between Jan. 2004 and Dec. 2015 – regardless of ART exposure duration, ART adherence, and patient’s status, were enrolled. ART-naïve, and patients <15 years old were excluded.

### Sample Size and Sampling Procedure

Study’s sample size and sampling procedure have also been [explained in another paper](https://www.medrxiv.org/content/10.1101/2023.11.14.23298518v1). Sample size was determined using double population proportion procedure recommended for prevalence studies [11-16] using the following equation:

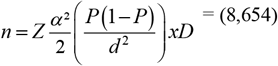

*Where: n = is the sample size*,

*P = Prevalence of opportunistic disease in PLHIV in Malawi @10*.*6% HIV rate based on 2010 MDHS*

*d = margin of error between sample and population sizes (n vs N) (0*.*006)*

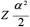*= statistical critical value at 95% confidence interval (1*.*96)*

*D = design effect for a cluster sampling of sampling frames (DHOs, Health facilities & ART Master cards) estimated @ 2*.*0*.

To compensate for poor documentation, a 15% allowance was applied to sample size (n=8,654) resulting in final sample size of (n=9,953). A three-stage probability sampling method was used to randomise the primary sampling units. The study’s sampling units (data frames) were health facilities offering HIV treatment and HIV patients’ medical records (PMR). By 2015, Malawi had total of 716 health facilities offering ART [17, 18] from which participating health facilities were sampled from using probability proportional to size using the equation …. *K=N/n:*

> *…*where *…K* = is the sampling interval unit, …*N* = is the total cumulative population size (or the sampling frame (population), …*n* = is the number of sampling units to be selected [19-22].

At each research (ART) centre, exclusion criteria were applied to the primary data frame; (PMR) [(*N*_1_)], to form secondary sampling frame, [*N*_2_]; a stratum of eligible patients. The sampling procedure was [then] replicated to randomly sample potential participants from *N*_2_. De-identified patients’ serial numbers [from (*N* _2_)] were listed in excel as a final sampling frame from which *n*1,*n*2,*n*3,*n*4,*n*5,………*nx* serial numbers were randomly selected. In research sites where PMR storage systems were fully functional electronic platforms; from the constructed sampling frame of eligible subjects (*N*_2_), participants were randomly selected using computer-generated random sample in excel. The per-centre sub-sample size allocation was determined based on the proportion of total number of ART-exposed adults against total sample size.

### Data collection tools and procedures

Data were collected using a structured electronic tool purposefully designed and developed in Microsoft Access. The tool was designed to capture data on different parameters (variables) including HIV history, ART history, de-identified demographic details, patients outcomes, AIDS conditions, and associated risk factors. Following initial wrangling, data was migrated to Stata for further cleansing and analysis.

### Data Quality Assurance

At each site, a resident research team was elected and trained on data collection procedures. The teams strictly followed the planned and approved data collection process. Close supervision of research teams during data collection and random double data re-entry checks were implemented as part of quality control measures.

### Statistical analysis procedure

Stata for windows version 15.1 was used to analyse data. Survival analysis involved (nonparametric) estimations of time-to-death (TTD) survival function with Kaplan-Meier (K-M) methods. Frequency and proportions values were expressed as absolute numbers and percentages. A day was used as time scale to calculate time-to-death from HIV/AIDS. Person-days of observation was calculated as date of death (event-function date) subtract from ART-initiation date. Descriptive survival analysis such as life table and K-M function curves were computed to estimate survival probability. The K-M Life Table was used to check probabilities of TTD at different time intervals and to check cumulative survival probabilities. Survival function equality tests across different levels of variables (covariates) were tested using the log-rank test and survival (or hazard) function curves. Both univariate and multivariate Cox Proportional Hazard (PH) regression model analyses were performed to investigate potential predictors of TTD. For univariate model, variables were purposefully selected based on their clinical significance. For multivariate Cox PH model, the predictors with univariate *p-*value (*p<*0.25) and non-violation of proportionality test (*p>*0.05) were included into the final multivariate model. The K-M curves were used to explore shapes of categorical predictors and determine their proportionality and inclusion in final multivariate model. The predictors’ fitness for the model was numerically verified using the Log-rank tests for equality across strata… [*model*…*1*]. In addition, the final model was double fitted using stepwise backward elimination of variables to judge the fitness of the model… [*model…2*]. Overall, results from [*model…1*] and [*mode…2*] were similar in both the number of variables and the effect size (ES) estimates. The Akaike’s Information Criterion (AIC) was used to evaluate the strength and validity of parametric models and the model that showed smaller statistic value; i.e., lowest AIC value, was the best-fit model. The final Cox PH model was then compared with the Weibull model – which was the best of the parametric distribution models, through Cox-Snell residual reliability test to judge the goodness-of-fit of model best-fitting the data. The Schoenfeld and scaled Schoenfeld residuals test was employed to assess final hazard regression model adequacy and numeric results were confirmed through scaled Schoenfeld residual curves. Hazard Ratios with 95% confidence intervals (HR, 95%CI) was computed and statistical significance was declared at 5% level (*p*<0.05). Thus, the Cox proportional hazard regression predictive model was used to determine factors associated with risk of time-to-death by controlling confounding factors.

### Operational definitions

#### Baseline (initial) test results

defines test results ideally done prior to commencing treatment, e.g. baseline CD4 cell count, baseline viral load (VL) level or count.

#### Repeat (Recent, Subsequent) test results

defines most recent (or last recorded) lab test results in treatment-experienced patients, e.g., recent CD4+ count, recent /subsequent VL level.

## RESULTS

### Baseline socio-demographic characteristics

The study’s sample disposition is presented in Table 1. Data for (*n*=9,953) enrolled patients were generated over an 11-year period – spanning from 2004 to 2015. Most patients were females (60.8%), young adults of 20-39 years (45.2%), and 78.8% were married. 33.7% & 32.8% had only basic education and secondary education levels respectively, and 55.3% were in full-time paid employment. 59.6% were from Southern region of Malawi, 67.7% from tertiary (3^rd^ tier) Health facilities (i.e., Central hospitals ART centres) and 65.1%, (N=6,477) were urban residents.

**Table 1:** Life Table of Time to Death Period Survival Function in PLHIV, Malawi, 2004-2015.

| Interval (days) | Number at risk (N<sub>i</sub>) | Deaths (failure) (D<sub>i</sub>) | Lost (censored) (C<sub>i</sub>) | Survival [probability] (S<sub>i</sub>) | Cum. Failure (1-S<sub>i</sub>) | Hazard | SE of estimated survival (S<sub>i</sub>) | [95% CI] |
| --- | --- | --- | --- | --- | --- | --- | --- | --- |
| 1 9 | 9953 | 0 | 13 | 1.0000 | 0.0000 | 0.0000 | 0.0000 | . |
| 9 29 | 9940 | 1 | 23 | 0.9999 | 0.0001 | 0.0000 | 0.0001 | 0.9993–0.9999 |
| 29 34 | 9916 | 1 | 7 | 0.9998 | 0.0002 | 0.0000 | 0.0001 | 0.9992–0.9999 |
| 34 36 | 9908 | 1 | 3 | 0.9997 | 0.0003 | 0.0001 | 0.0002 | 0.9991–0.9999 |
| 36 37 | 9904 | 3 | 0 | 0.9994 | 0.0006 | 0.0003 | 0.0002 | 0.9987–0.9997 |
| 37 40 | 9901 | 1 | 4 | 0.9993 | 0.0007 | 0.0000 | 0.0003 | 0.9985–0.9997 |
| 40 41 | 9896 | 1 | 2 | 0.9992 | 0.0008 | 0.0001 | 0.0003 | 0.9984–0.9996 |
| 41 42 | 9893 | 1 | 3 | 0.9991 | 0.0009 | 0.0001 | 0.0003 | 0.9983–0.9995 |
| 42 43 | 9889 | 1 | 1 | 0.9990 | 0.0010 | 0.0001 | 0.0003 | 0.9981–0.9995 |
| 43 52 | 9887 | 2 | 7 | 0.9988 | 0.0012 | 0.0000 | 0.0003 | 0.9979–0.9993 |
| 52 54 | 9878 | 1 | 6 | 0.9987 | 0.0013 | 0.0001 | 0.0004 | 0.9977–0.9992 |
| 54 156 | 9871 | 34 | 147 | 0.9952 | 0.0048 | 0.0000 | 0.0007 | 0.9936–0.9964 |
| 156 158 | 9690 | 2 | 3 | 0.9950 | 0.0050 | 0.0001 | 0.0007 | 0.9934–0.9962 |
| 158 250 | 9685 | 27 | 129 | 0.9922 | 0.0078 | 0.0000 | 0.0009 | 0.9903–0.9938 |
| 250 500 | 9529 | 90 | 379 | 0.9827 | 0.0173 | 0.0000 | 0.0013 | 0.9798–0.9851 |
| 500 750 | 9060 | 107 | 379 | 0.9708 | 0.0292 | 0.0000 | 0.0017 | 0.9672–0.9740 |
| 750 1000 | 8574 | 92 | 449 | 0.9601 | 0.0399 | 0.0000 | 0.0020 | 0.9559–0.9639 |
| 1000 1250 | 8033 | 100 | 501 | 0.9478 | 0.0522 | 0.0001 | 0.0024 | 0.9429–0.9522 |
| 1250 1500 | 7432 | 89 | 519 | 0.9360 | 0.0640 | 0.0000 | 0.0026 | 0.9306–0.9410 |
| 1500 1750 | 6824 | 87 | 483 | 0.9236 | 0.0764 | 0.0001 | 0.0029 | 0.9177–0.9292 |
| 1750 2000 | 6254 | 91 | 463 | 0.9097 | 0.0903 | 0.0001 | 0.0032 | 0.9032–0.9158 |
| 2000 2500 | 5700 | 137 | 895 | 0.8860 | 0.1140 | 0.0001 | 0.0037 | 0.8784–0.8930 |
| 2500 3000 | 4668 | 99 | 1088 | 0.8647 | 0.1353 | 0.0000 | 0.0042 | 0.8562–0.8727 |
| 3000 4000 | 3481 | 126 | 2091 | 0.8200 | 0.1800 | 0.0001 | 0.0056 | 0.8088–0.8306 |
| 4000 5000 | 1264 | 58 | 601 | 0.7706 | 0.2294 | 0.0001 | 0.0082 | 0.7541–0.7862 |
| 5000 7000 | 605 | 23 | 536 | 0.7180 | 0.2820 | 0.0000 | 0.0130 | 0.6915–0.7427 |
| 7000 9000 | 46 | 2 | 40 | 0.6628 | 0.3372 | 0.0000 | 0.0394 | 0.5792–0.7336 |
| 9000 9161 | 4 | 0 | 1 | 0.6628 | 0.3372 | 0.0000 | 0.0394 | 0.5792–0.7336 |
| 9161 9172 | 3 | 0 | 1 | 0.6628 | 0.3372 | 0.0000 | 0.0394 | 0.5792–0.7336 |
| 9172 9224 | 2 | 0 | 1 | 0.6628 | 0.3372 | 0.0000 | 0.0394 | 0.5792–0.7336 |
| 9224 9250 | 1 | 0 | 1 | 0.6628 | 0.3372 | 0.0000 | 0.0394 | 0.5792–0.7336 |

**Key:** N<sub>i</sub> = number of participants who are event free and considered at risk during interval t D<sub>i</sub> = number of participants who die (or suffer the event of interest) during interval t C<sub>i</sub> = number of participants who are censored during interval t S<sub>i</sub> = the proportion surviving (or remaining event free) past interval t; (also called the cumulative survival probability) 1-S<sub>i</sub> = Cumulative incidence (or cumulative failure probability)

All patients were treatment-experienced. 48.1% and 24.5% of patients had WHO clinical stage III & IV manifestations when they commenced treatment and 27.5% were asymptomatic; of which 24.9% of patients had low CD4 count and 2.6% commenced ART under PMTCT’s Option-B+ criteria (pregnant or breastfeeding mothers).

The mean (±SD) baseline CD4 count was 145.1±95.1 cells/µL. 69.9% of patients were severely immunosuppressed with baseline CD4 count <250 cells/µL and correspondingly severe viral failure of VL>1,000 copies/mL (>3.0 Log_10_) was present in 96.4% of patients. The mean (±SD) baseline VL was 6.0±4.3 Log_10_ copies/mL. 86.5% had lived with HIV for over 2 years, 78.4% had been on ART more than 2 years and 67.9% were continuing treatment with good adherence rate. 67.1% of patients were commenced on Efavirenz-based Reverse Transcriptase Inhibitors ART as first-line treatment regimens compared to 28.5% on NVP-based regimens. 95.4% were continuing with first-line regimens while 3.3% had been switched to second-line regimens; the boosted-Ritonavir PIs regimens – implying that they had proven first-line treatment failure.

Death occurred more in female patients (58.7%) compared to 41.7% in males and was higher among 40-54 year olds (39.8%) but least among 15-19 years olds (2.1%). More deaths had also occurred among patients from Thyolo District Hospital ART centre; a semi-urban secondary level health facility in Southern region (32.8%) followed by those from Mzuzu Central hospital ART centre; an urban tertiary level health facility in Northern Region (26.3%), but was least in patients from Dream Programmes ART Centre (2.1%) whereas Nkhoma and PIH hospitals ART Centres recorded nil deaths. Correspondingly, tertiary level health facilities registered an overall 60.1% mortality compared to 2.1% from primary level facilities. 61.4% and 63.1% of deaths occurred in urban areas and in Southern region of Malawi, respectively. Mycobacterial conditions were responsible for 38.1% mortality but was least in neoplastic conditions (2.1%), and patients who were asymptomatic when treatment was commenced registered highest mortality (31.8%) among the clinical presentation predictor’s classes. However, in parallel to aetiological causes, patients with advanced HIV disease; WHO stage III, registered the highest mortality rate of 56.1% compared to 1.9% among PMTCT’s Option-B+. Expectedly, low CD4 count and high VL were greatest predictors of death. Thus., recent (repeat) CD4 count <500 cells/µL was responsible for 91.2% mortality compared to 0.4% in >1000 cells/µL CD4 count, and correspondingly, recent VL>1000 copies/mL was responsible for 93.5% mortality compared to 6.5% in VL 40-1000 copies/mL while those with undetectable VL (<40c/mL) registered nil (0.0%) deaths.

### Survival Times-to-Death Findings

Descriptive analysis findings of time-to-death survival function involved a sub-cohort of (N=1,177) patients who died within the 11-year period data was generated (2004-2015) from the total (n=9,953) enrolled subjects. The findings show that all patients had single entry into the study. Exit time ranged from 1 to 9,224 days with the mean value of 2,421.9 days, occurring at the rate of 0.00004883 event-failure per-person-day.

The Kaplan-Meier Actuarial Life table of TTD survival function (Table 2) is drawn to different time-interval bandwidths scales of 250, 500, 1000, and 2000 days. The top 14 and the bottom 4 rows are precise time-intervals (days) on which the failure-event (death) occurred and the total daily mortality. Accordingly, the study design allowed no left censoring (i.e., no deaths before ART-initiation were accounted for). 76 deaths occurred within the first time-interval band: between 1 and 250 time-interval days (8.2 months). Precisely, the first and second deaths (failure-events) occurred between 9 and 29, and between 29 and 34 time-intervals respectively. 11.6% of all deaths (N=137), occurred between 2000 and 2500 time-interval days, followed by (N=126) deaths between 3000 and 4000 time-interval. Two female patients aged 35 and 67 years; both from Lighthouse Trust ART centre, a tertiary facility at KCH, died day-1 after commencing treatment.

**Table 2:**
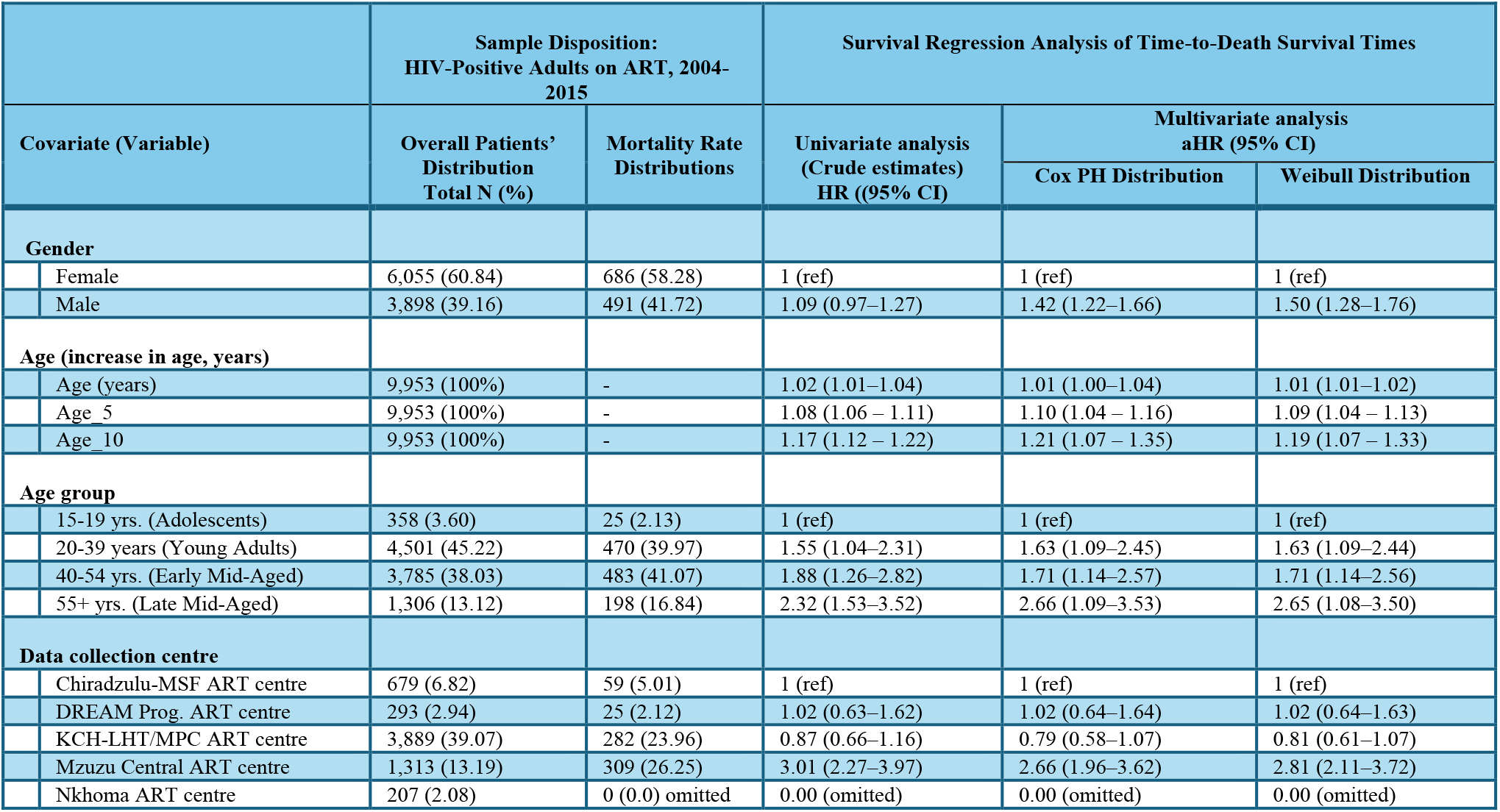
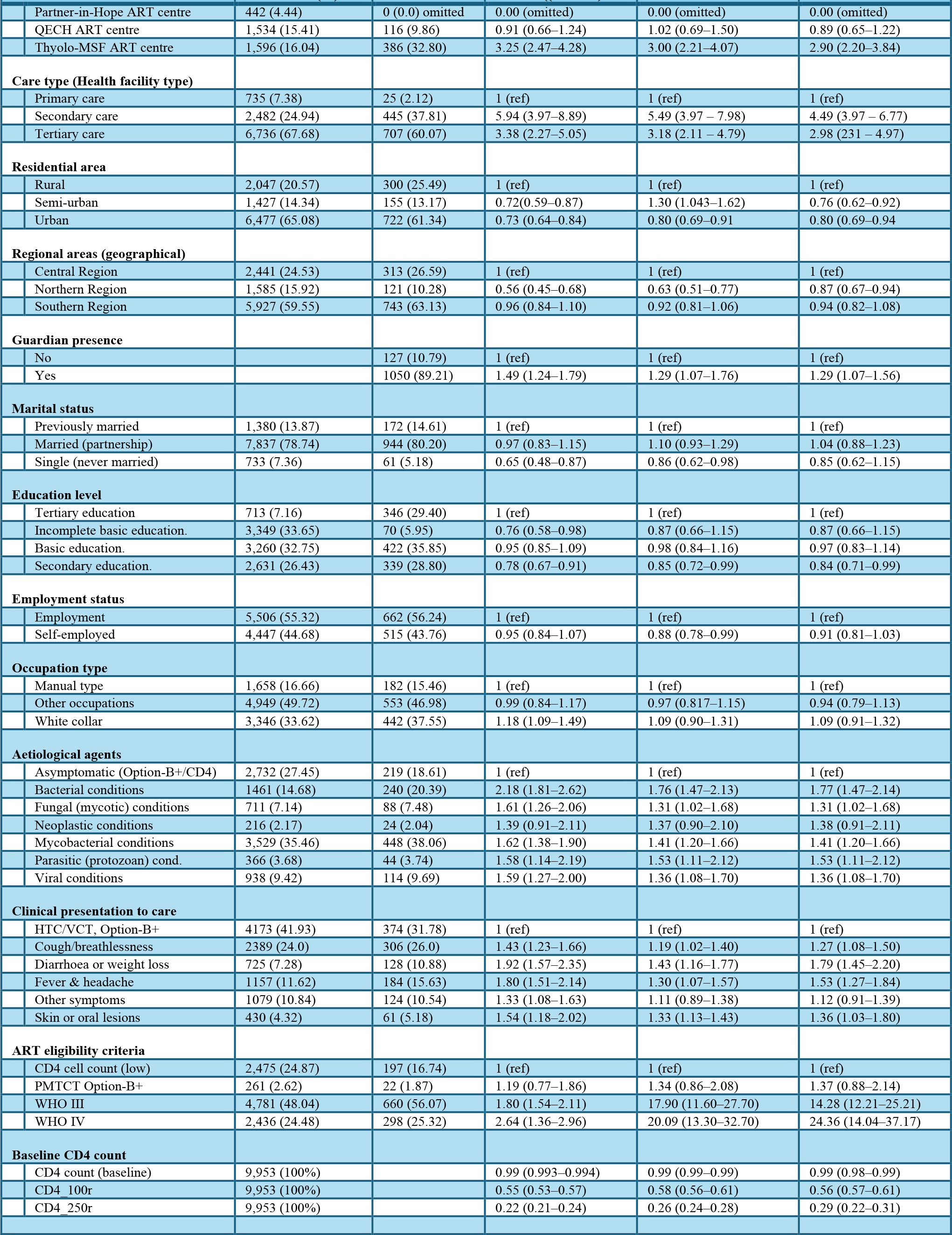
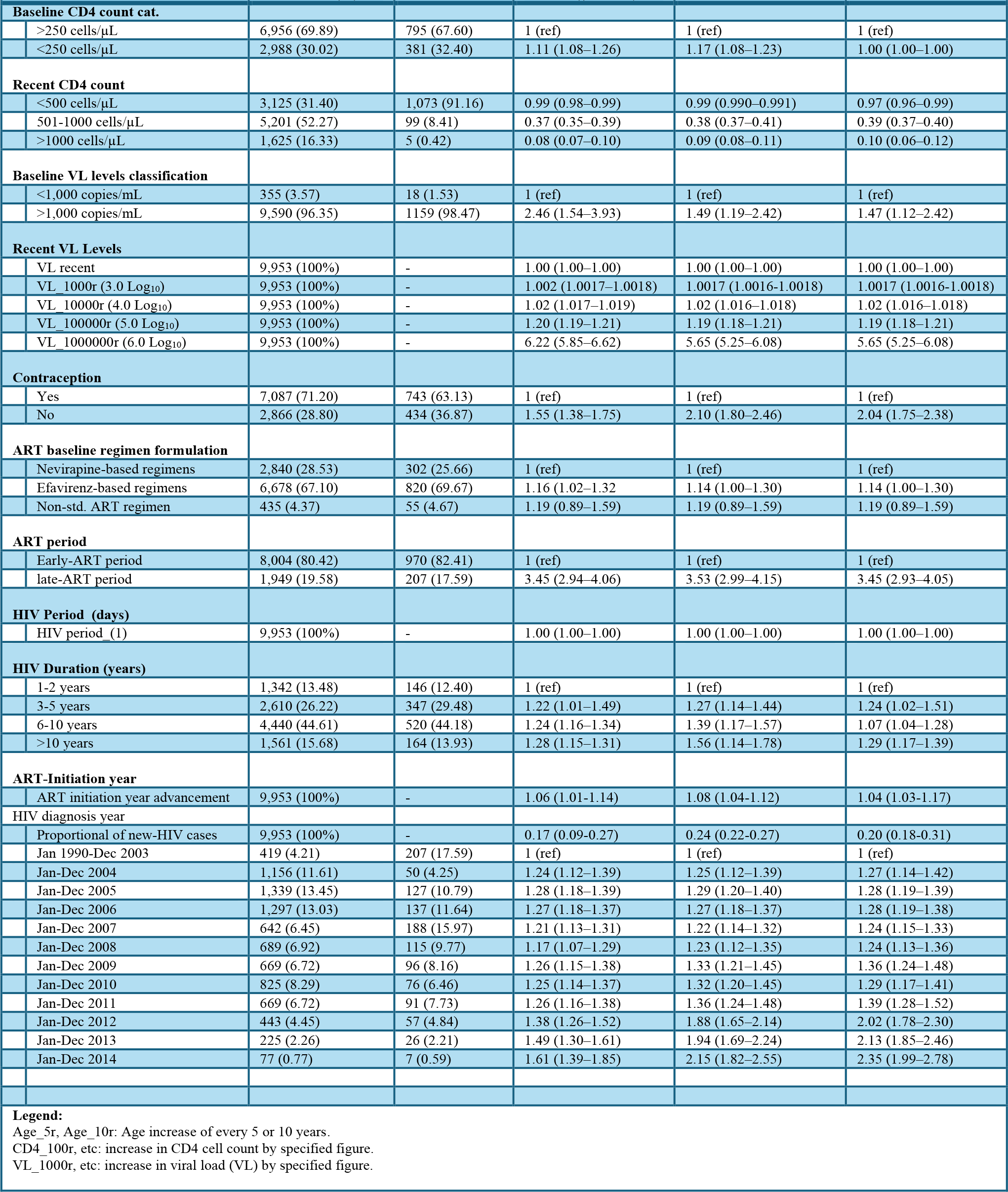
Univariate and Multivariate Survival Regression of Time-to-Death Survival Function.

|  |  | Sample Disposition:<br>HIV-Positive Adults on ART, 2004-2015 |  | Survival Regression Analysis of Time-to-Death Survival Times |  |  |
| --- | --- | --- | --- | --- | --- | --- |
| Covariate (Variable) |  | Overall Patients' Distribution<br>Total N (%) | Mortality Rate Distributions | Univariate analysis<br>(Crude estimates)<br>HR ((95% CI) | Multivariate analysis<br>aHR (95% CI) |  |
|  |  |  |  |  | Cox PH Distribution | Weibull Distribution |
| Gender |  |  |  |  |  |  |
|  | Female | 6,055 (60.84) | 686 (58.28) | 1 (ref) | 1 (ref) | 1 (ref) |
|  | Male | 3,898 (39.16) | 491 (41.72) | 1.09 (0.97–1.27) | 1.42 (1.22–1.66) | 1.50 (1.28–1.76) |
| Age (increase in age, years) |  |  |  |  |  |  |
|  | Age (years) | 9,953 (100%) | - | 1.02 (1.01–1.04) | 1.01 (1.00–1.04) | 1.01 (1.01–1.02) |
|  | Age_5 | 9,953 (100%) | - | 1.08 (1.06 – 1.11) | 1.10 (1.04 – 1.16) | 1.09 (1.04 – 1.13) |
|  | Age_10 | 9,953 (100%) | - | 1.17 (1.12 – 1.22) | 1.21 (1.07 – 1.35) | 1.19 (1.07 – 1.33) |
| Age group |  |  |  |  |  |  |
|  | 15-19 yrs. (Adolescents) | 358 (3.60) | 25 (2.13) | 1 (ref) | 1 (ref) | 1 (ref) |
|  | 20-39 years (Young Adults) | 4,501 (45.22) | 470 (39.97) | 1.55 (1.04–2.31) | 1.63 (1.09–2.45) | 1.63 (1.09–2.44) |
|  | 40-54 yrs. (Early Mid-Aged) | 3,785 (38.03) | 483 (41.07) | 1.88 (1.26–2.82) | 1.71 (1.14–2.57) | 1.71 (1.14–2.56) |
|  | 55+ yrs. (Late Mid-Aged) | 1,306 (13.12) | 198 (16.84) | 2.32 (1.53–3.52) | 2.66 (1.09–3.53) | 2.65 (1.08–3.50) |
| Data collection centre |  |  |  |  |  |  |
|  | Chiradzulu-MSF ART centre | 679 (6.82) | 59 (5.01) | 1 (ref) | 1 (ref) | 1 (ref) |
|  | DREAM Prog. ART centre | 293 (2.94) | 25 (2.12) | 1.02 (0.63–1.62) | 1.02 (0.64–1.64) | 1.02 (0.64–1.63) |
|  | KCH-LHT/MPC ART centre | 3,889 (39.07) | 282 (23.96) | 0.87 (0.66–1.16) | 0.79 (0.58–1.07) | 0.81 (0.61–1.07) |
|  | Mzuzu Central ART centre | 1,313 (13.19) | 309 (26.25) | 3.01 (2.27–3.97) | 2.66 (1.96–3.62) | 2.81 (2.11–3.72) |
|  | Nkhoma ART centre | 207 (2.08) | 0 (0.0) omitted | 0.00 (omitted) | 0.00 (omitted) | 0.00 (omitted) |

|  |  | Sample Disposition:<br>HIV-Positive Adults on ART, 2004-2015 | Survival Regression Analysis of Time-to-Death Survival Times |  |  |  |
| --- | --- | --- | --- | --- | --- | --- |
| Covariate (Variable) |  | Overall Patients' Distribution<br>Total N (%) | Mortality Rate Distributions | Univariate analysis<br>(Crude estimates)<br>HR (95% CI) | Multivariate analysis<br>aHR (95% CI) |  |
|  |  |  |  |  | Cox PH Distribution | Weibull Distribution |
|  | Partner-in-Hope ART centre | 442 (4.44) | 0 (0.0) omitted | 0.00 (omitted) | 0.00 (omitted) | 0.00 (omitted) |
|  | QECH ART centre | 1,534 (15.41) | 116 (9.86) | 0.91 (0.66–1.24) | 1.02 (0.69–1.50) | 0.89 (0.65–1.22) |
|  | Thyolo-MSF ART centre | 1,596 (16.04) | 386 (32.80) | 3.25 (2.47–4.28) | 3.00 (2.21–4.07) | 2.90 (2.20–3.84) |
| Care type (Health facility type) |  |  |  |  |  |  |
|  | Primary care | 735 (7.38) | 25 (2.12) | 1 (ref) | 1 (ref) | 1 (ref) |
|  | Secondary care | 2,482 (24.94) | 445 (37.81) | 5.94 (3.97–8.89) | 5.49 (3.97 – 7.98) | 4.49 (3.97 – 6.77) |
|  | Tertiary care | 6,736 (67.68) | 707 (60.07) | 3.38 (2.27–5.05) | 3.18 (2.11 – 4.79) | 2.98 (231 – 4.97) |
| Residential area |  |  |  |  |  |  |
|  | Rural | 2,047 (20.57) | 300 (25.49) | 1 (ref) | 1 (ref) | 1 (ref) |
|  | Semi-urban | 1,427 (14.34) | 155 (13.17) | 0.72(0.59–0.87) | 1.30 (1.043–1.62) | 0.76 (0.62–0.92) |
|  | Urban | 6,477 (65.08) | 722 (61.34) | 0.73 (0.64–0.84) | 0.80 (0.69–0.91) | 0.80 (0.69–0.94) |
| Regional areas (geographical) |  |  |  |  |  |  |
|  | Central Region | 2,441 (24.53) | 313 (26.59) | 1 (ref) | 1 (ref) | 1 (ref) |
|  | Northern Region | 1,585 (15.92) | 121 (10.28) | 0.56 (0.45–0.68) | 0.63 (0.51–0.77) | 0.87 (0.67–0.94) |
|  | Southern Region | 5,927 (59.55) | 743 (63.13) | 0.96 (0.84–1.10) | 0.92 (0.81–1.06) | 0.94 (0.82–1.08) |
| Guardian presence |  |  |  |  |  |  |
|  | No |  | 127 (10.79) | 1 (ref) | 1 (ref) | 1 (ref) |
|  | Yes |  | 1050 (89.21) | 1.49 (1.24–1.79) | 1.29 (1.07–1.76) | 1.29 (1.07–1.56) |
| Marital status |  |  |  |  |  |  |
|  | Previously married | 1,380 (13.87) | 172 (14.61) | 1 (ref) | 1 (ref) | 1 (ref) |
|  | Married (partnership) | 7,837 (78.74) | 944 (80.20) | 0.97 (0.83–1.15) | 1.10 (0.93–1.29) | 1.04 (0.88–1.23) |
|  | Single (never married) | 733 (7.36) | 61 (5.18) | 0.65 (0.48–0.87) | 0.86 (0.62–0.98) | 0.85 (0.62–1.15) |
| Education level |  |  |  |  |  |  |
|  | Tertiary education | 713 (7.16) | 346 (29.40) | 1 (ref) | 1 (ref) | 1 (ref) |
|  | Incomplete basic education. | 3,349 (33.65) | 70 (5.95) | 0.76 (0.58–0.98) | 0.87 (0.66–1.15) | 0.87 (0.66–1.15) |
|  | Basic education. | 3,260 (32.75) | 422 (35.85) | 0.95 (0.85–1.09) | 0.98 (0.84–1.16) | 0.97 (0.83–1.14) |
|  | Secondary education. | 2,631 (26.43) | 339 (28.80) | 0.78 (0.67–0.91) | 0.85 (0.72–0.99) | 0.84 (0.71–0.99) |
| Employment status |  |  |  |  |  |  |
|  | Employment | 5,506 (55.32) | 662 (56.24) | 1 (ref) | 1 (ref) | 1 (ref) |
|  | Self-employed | 4,447 (44.68) | 515 (43.76) | 0.95 (0.84–1.07) | 0.88 (0.78–0.99) | 0.91 (0.81–1.03) |
| Occupation type |  |  |  |  |  |  |
|  | Manual type | 1,658 (16.66) | 182 (15.46) | 1 (ref) | 1 (ref) | 1 (ref) |
|  | Other occupations | 4,949 (49.72) | 553 (46.98) | 0.99 (0.84–1.17) | 0.97 (0.817–1.15) | 0.94 (0.79–1.13) |
|  | White collar | 3,346 (33.62) | 442 (37.55) | 1.18 (1.09–1.49) | 1.09 (0.90–1.31) | 1.09 (0.91–1.32) |
| Aetiological agents |  |  |  |  |  |  |
|  | Asymptomatic (Option-B+/CD4) | 2,732 (27.45) | 219 (18.61) | 1 (ref) | 1 (ref) | 1 (ref) |
|  | Bacterial conditions | 1461 (14.68) | 240 (20.39) | 2.18 (1.81–2.62) | 1.76 (1.47–2.13) | 1.77 (1.47–2.14) |
|  | Fungal (mycotic) conditions | 711 (7.14) | 88 (7.48) | 1.61 (1.26–2.06) | 1.31 (1.02–1.68) | 1.31 (1.02–1.68) |
|  | Neoplastic conditions | 216 (2.17) | 24 (2.04) | 1.39 (0.91–2.11) | 1.37 (0.90–2.10) | 1.38 (0.91–2.11) |
|  | Mycobacterial conditions | 3,529 (35.46) | 448 (38.06) | 1.62 (1.38–1.90) | 1.41 (1.20–1.66) | 1.41 (1.20–1.66) |
|  | Parasitic (protozoan) cond. | 366 (3.68) | 44 (3.74) | 1.58 (1.14–2.19) | 1.53 (1.11–2.12) | 1.53 (1.11–2.12) |
|  | Viral conditions | 938 (9.42) | 114 (9.69) | 1.59 (1.27–2.00) | 1.36 (1.08–1.70) | 1.36 (1.08–1.70) |
| Clinical presentation to care |  |  |  |  |  |  |
|  | HTC/VCT, Option-B+ | 4173 (41.93) | 374 (31.78) | 1 (ref) | 1 (ref) | 1 (ref) |
|  | Cough/shortness of breath | 2389 (24.0) | 306 (26.0) | 1.43 (1.23–1.66) | 1.19 (1.02–1.40) | 1.27 (1.08–1.50) |
|  | Diarrhoea or weight loss | 725 (7.28) | 128 (10.88) | 1.92 (1.57–2.35) | 1.43 (1.16–1.77) | 1.79 (1.45–2.20) |
|  | Fever & headache | 1157 (11.62) | 184 (15.63) | 1.80 (1.51–2.14) | 1.30 (1.07–1.57) | 1.53 (1.27–1.84) |
|  | Other symptoms | 1079 (10.84) | 124 (10.54) | 1.33 (1.08–1.63) | 1.11 (0.89–1.38) | 1.12 (0.91–1.39) |
|  | Skin or oral lesions | 430 (4.32) | 61 (5.18) | 1.54 (1.18–2.02) | 1.33 (1.13–1.43) | 1.36 (1.03–1.80) |
| ART eligibility criteria |  |  |  |  |  |  |
|  | CD4 cell count (low) | 2,475 (24.87) | 197 (16.74) | 1 (ref) | 1 (ref) | 1 (ref) |
|  | PMTCT Option-B+ | 261 (2.62) | 22 (1.87) | 1.19 (0.77–1.86) | 1.34 (0.86–2.08) | 1.37 (0.88–2.14) |
|  | WHO III | 4,781 (48.04) | 660 (56.07) | 1.80 (1.54–2.11) | 17.90 (11.60–27.70) | 14.28 (12.21–25.21) |
|  | WHO IV | 2,436 (24.48) | 298 (25.32) | 2.64 (1.36–2.96) | 20.09 (13.30–32.70) | 24.36 (14.04–37.17) |
| Baseline CD4 count |  |  |  |  |  |  |
|  | CD4 count (baseline) | 9,953 (100%) |  | 0.99 (0.993–0.994) | 0.99 (0.99–0.99) | 0.99 (0.98–0.99) |
|  | CD4_100r | 9,953 (100%) |  | 0.55 (0.53–0.57) | 0.58 (0.56–0.61) | 0.56 (0.57–0.61) |
|  | CD4_250r | 9,953 (100%) |  | 0.22 (0.21–0.24) | 0.26 (0.24–0.28) | 0.29 (0.22–0.31) |
| Covariate (Variable) |  | Overall Patients' Distribution<br>Total N (%) | Mortality Rate Distributions | Univariate analysis<br>(Crude estimates)<br>HR (95% CI) | Multivariate analysis<br>aHR (95% CI) |  |
|  |  |  |  |  | Cox PH Distribution | Weibull Distribution |
| Baseline CD4 count cat. |  |  |  |  |  |  |
|  | >250 cells/μL | 6,956 (69.89) | 795 (67.60) | 1 (ref) | 1 (ref) | 1 (ref) |
|  | <250 cells/μL | 2,988 (30.02) | 381 (32.40) | 1.11 (1.08–1.26) | 1.17 (1.08–1.23) | 1.00 (1.00–1.00) |
| Recent CD4 count |  |  |  |  |  |  |
|  | <500 cells/μL | 3,125 (31.40) | 1,073 (91.16) | 0.99 (0.98–0.99) | 0.99 (0.990–0.991) | 0.97 (0.96–0.99) |
|  | 501-1000 cells/μL | 5,201 (52.27) | 99 (8.41) | 0.37 (0.35–0.39) | 0.38 (0.37–0.41) | 0.39 (0.37–0.40) |
|  | >1000 cells/μL | 1,625 (16.33) | 5 (0.42) | 0.08 (0.07–0.10) | 0.09 (0.08–0.11) | 0.10 (0.06–0.12) |
| Baseline VL levels classification |  |  |  |  |  |  |
|  | <1,000 copies/mL | 355 (3.57) | 18 (1.53) | 1 (ref) | 1 (ref) | 1 (ref) |
|  | >1,000 copies/mL | 9,590 (96.35) | 1159 (98.47) | 2.46 (1.54–3.93) | 1.49 (1.19–2.42) | 1.47 (1.12–2.42) |
| Recent VL Levels |  |  |  |  |  |  |
|  | VL recent | 9,953 (100%) | - | 1.00 (1.00–1.00) | 1.00 (1.00–1.00) | 1.00 (1.00–1.00) |
|  | VL_1000r (3.0 Log<sub>10</sub>) | 9,953 (100%) | - | 1.002 (1.0017–1.0018) | 1.0017 (1.0016-1.0018) | 1.0017 (1.0016-1.0018) |
|  | VL_10000r (4.0 Log<sub>10</sub>) | 9,953 (100%) | - | 1.02 (1.017–1.019) | 1.02 (1.016–1.018) | 1.02 (1.016–1.018) |
|  | VL_100000r (5.0 Log<sub>10</sub>) | 9,953 (100%) | - | 1.20 (1.19–1.21) | 1.19 (1.18–1.21) | 1.19 (1.18–1.21) |
|  | VL_1000000r (6.0 Log<sub>10</sub>) | 9,953 (100%) | - | 6.22 (5.85–6.62) | 5.65 (5.25–6.08) | 5.65 (5.25–6.08) |
| Contraception |  |  |  |  |  |  |
|  | Yes | 7,087 (71.20) | 743 (63.13) | 1 (ref) | 1 (ref) | 1 (ref) |
|  | No | 2,866 (28.80) | 434 (36.87) | 1.55 (1.38–1.75) | 2.10 (1.80–2.46) | 2.04 (1.75–2.38) |
| ART baseline regimen formulation |  |  |  |  |  |  |
|  | Nevirapine-based regimens | 2,840 (28.53) | 302 (25.66) | 1 (ref) | 1 (ref) | 1 (ref) |
|  | Efavirenz-based regimens | 6,678 (67.10) | 820 (69.67) | 1.16 (1.02–1.32) | 1.14 (1.00–1.30) | 1.14 (1.00–1.30) |
|  | Non-std. ART regimen | 435 (4.37) | 55 (4.67) | 1.19 (0.89–1.59) | 1.19 (0.89–1.59) | 1.19 (0.89–1.59) |
| ART period |  |  |  |  |  |  |
|  | Early-ART period | 8,004 (80.42) | 970 (82.41) | 1 (ref) | 1 (ref) | 1 (ref) |
|  | late-ART period | 1,949 (19.58) | 207 (17.59) | 3.45 (2.94–4.06) | 3.53 (2.99–4.15) | 3.45 (2.93–4.05) |
| HIV Period (days) |  |  |  |  |  |  |
|  | HIV period_ (1) | 9,953 (100%) | - | 1.00 (1.00–1.00) | 1.00 (1.00–1.00) | 1.00 (1.00–1.00) |
| HIV Duration (years) |  |  |  |  |  |  |
|  | 1-2 years | 1,342 (13.48) | 146 (12.40) | 1 (ref) | 1 (ref) | 1 (ref) |
|  | 3-5 years | 2,610 (26.22) | 347 (29.48) | 1.22 (1.01–1.49) | 1.27 (1.14–1.44) | 1.24 (1.02–1.51) |
|  | 6-10 years | 4,440 (44.61) | 520 (44.18) | 1.24 (1.16–1.34) | 1.39 (1.17–1.57) | 1.07 (1.04–1.28) |
|  | >10 years | 1,561 (15.68) | 164 (13.93) | 1.28 (1.15–1.31) | 1.56 (1.14–1.78) | 1.29 (1.17–1.39) |
| ART-Initiation year |  |  |  |  |  |  |
|  | ART initiation year advancement | 9,953 (100%) | - | 1.06 (1.01-1.14) | 1.08 (1.04-1.12) | 1.04 (1.03-1.17) |
| HIV diagnosis year |  |  |  |  |  |  |
|  | Proportional of new-HIV cases | 9,953 (100%) | - | 0.17 (0.09-0.27) | 0.24 (0.22-0.27) | 0.20 (0.18-0.31) |
|  | Jan 1990-Dec 2003 | 419 (4.21) | 207 (17.59) | 1 (ref) | 1 (ref) | 1 (ref) |
|  | Jan-Dec 2004 | 1,156 (11.61) | 50 (4.25) | 1.24 (1.12–1.39) | 1.25 (1.12–1.39) | 1.27 (1.14–1.42) |
|  | Jan-Dec 2005 | 1,339 (13.45) | 127 (10.79) | 1.28 (1.18–1.39) | 1.29 (1.20–1.40) | 1.28 (1.19–1.39) |
|  | Jan-Dec 2006 | 1,297 (13.03) | 137 (11.64) | 1.27 (1.18–1.37) | 1.27 (1.18–1.37) | 1.28 (1.19–1.38) |
|  | Jan-Dec 2007 | 642 (6.45) | 188 (15.97) | 1.21 (1.13–1.31) | 1.22 (1.14–1.32) | 1.24 (1.15–1.33) |
|  | Jan-Dec 2008 | 689 (6.92) | 115 (9.77) | 1.17 (1.07–1.29) | 1.23 (1.12–1.35) | 1.24 (1.13–1.36) |
|  | Jan-Dec 2009 | 669 (6.72) | 96 (8.16) | 1.26 (1.15–1.38) | 1.33 (1.21–1.45) | 1.36 (1.24–1.48) |
|  | Jan-Dec 2010 | 825 (8.29) | 76 (6.46) | 1.25 (1.14–1.37) | 1.32 (1.20–1.45) | 1.29 (1.17–1.41) |
|  | Jan-Dec 2011 | 669 (6.72) | 91 (7.73) | 1.26 (1.16–1.38) | 1.36 (1.24–1.48) | 1.39 (1.28–1.52) |
|  | Jan-Dec 2012 | 443 (4.45) | 57 (4.84) | 1.38 (1.26–1.52) | 1.88 (1.65–2.14) | 2.02 (1.78–2.30) |
|  | Jan-Dec 2013 | 225 (2.26) | 26 (2.21) | 1.49 (1.30–1.61) | 1.94 (1.69–2.24) | 2.13 (1.85–2.46) |
|  | Jan-Dec 2014 | 77 (0.77) | 7 (0.59) | 1.61 (1.39–1.85) | 2.15 (1.82–2.55) | 2.35 (1.99–2.78) |
| Legend:<br>Age_5r, Age_10r: Age increase of every 5 or 10 years.<br>CD4_100r, etc: increase in CD4 cell count by specified figure.<br>VL_1000r, etc: increase in viral load (VL) by specified figure. |  |  |  |  |  |  |

Figure 1 is a lifetable plot of TTD survival function showing overall and gender stratified curves. The graph shows unclear parallelism in gender stratified curves during the first 3000 analysis time-period followed by crossover at about 7100 analysis time period (Figure 1A). However, the plots showed that cumulative survival drops with every death until maximum death (N=1,177) was reached at 9,224 time-interval. This is also reflected in the overall Kaplan-Meier survival estimates, and the Nelson-Aalen cumulative hazard (mortality) estimate plots of TTD survival function (Figure 2) which demonstrate that the proportion of patients alive steadily declined over time, then suddenly dropped to flatten out (Figure 2A), while conversely, the curve for patients dying increased over time (Figure 2B).

**Figure 1:**
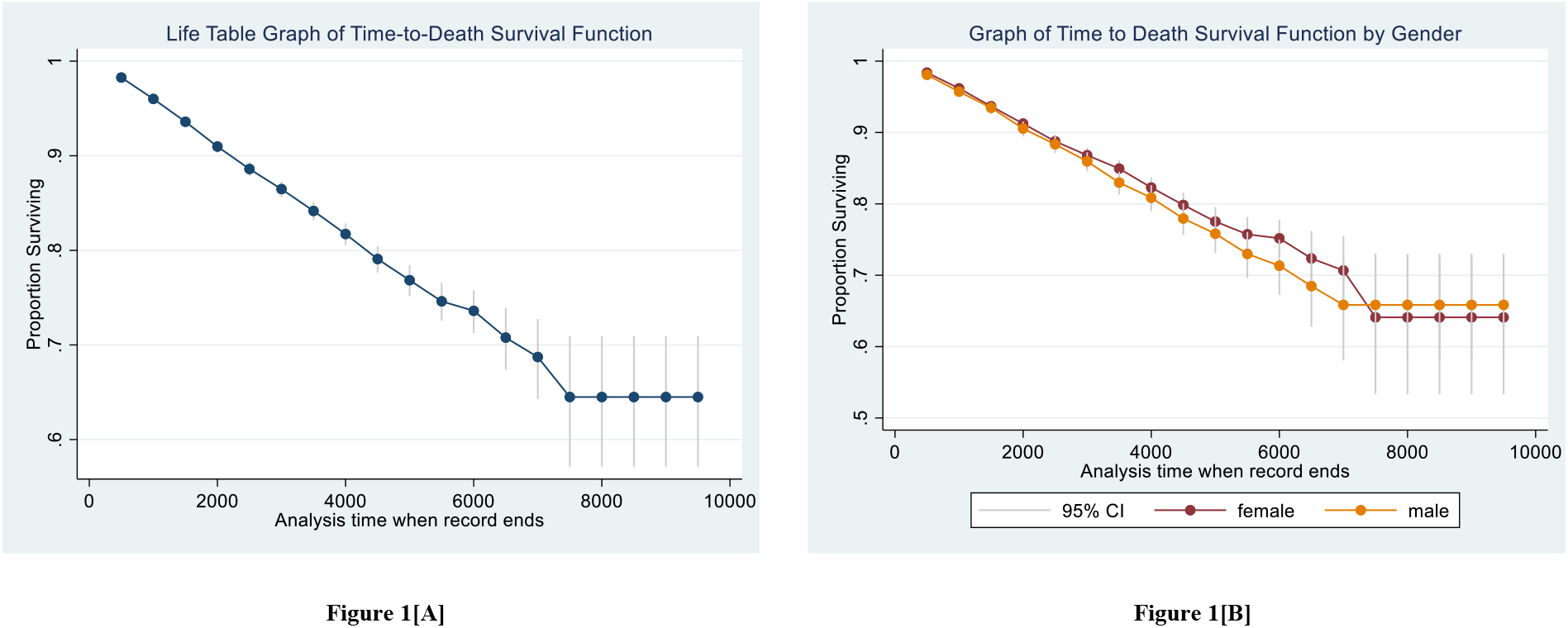
Lifetable Graphs of Time-to-Death Survival Function, Overall and by Gender. The gender stratified curve (Fig. 1B) shows somehow unclear parallelism during the first 3000^th^ analysis time-point with curve crossover about 7000^th^ analysis time-point. The vertical lines in the curves represent 95% CIs for the survivor function which are evidently widening as analysis time diminishes.

**Figure 2:**
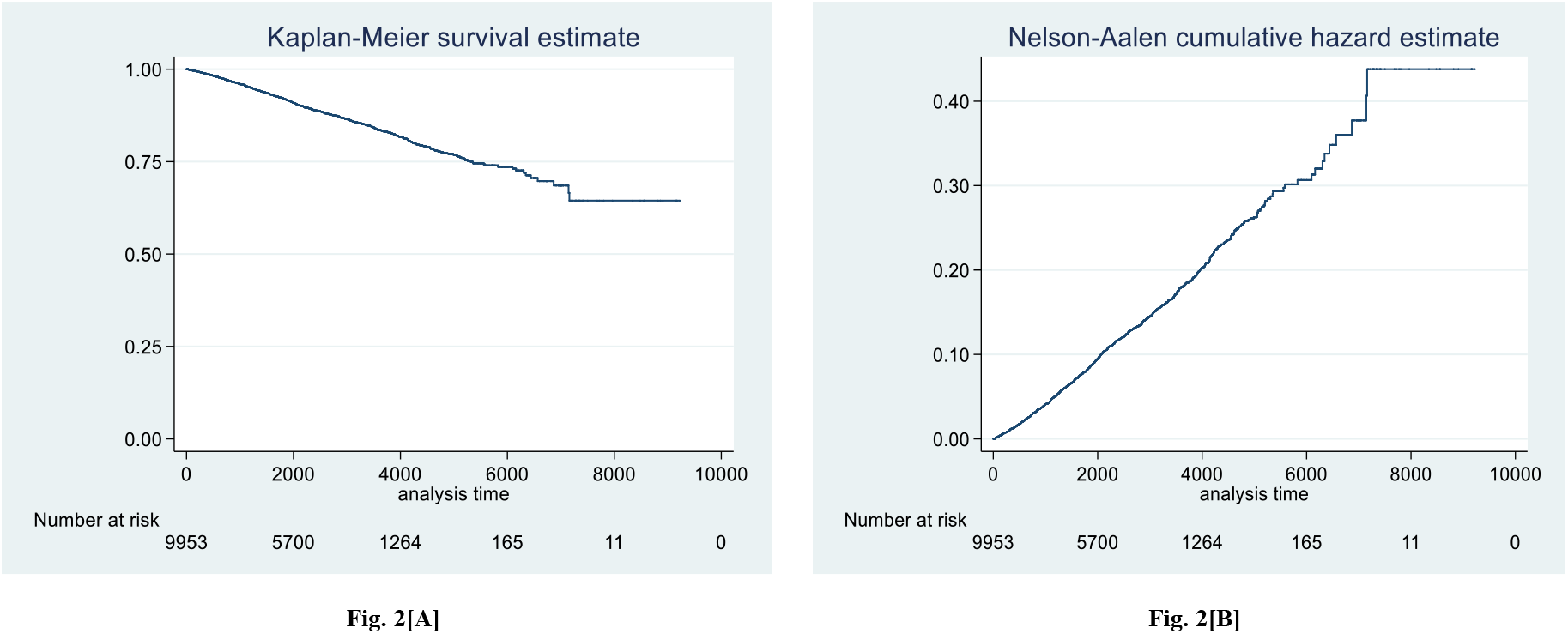
The K-M Survival Plot and the Nelson-Aalen Cummulative Hazard Estimate of TTD Survival Function. The Kaplan-Meier curve indicates a steadliy declining proportion (%) of patients surviving (alive) over time and a sudden drop and flattening out. The Nelson-Aalen conversely shows increasing hazard (of death) over time. Thus, the Cox and Weibull survival models were evaluated for best-fitness to the data by examining the shapes of their Cox-Snell plots and the appropriateness of the cumulative hazard rates to the reference line. The Cox PH model was judged best-fitting the data with best adherent model assumptions (cumulative hazard) closer to the diagonal reference line.

### Predictors Associated with Time-to-Death From HIV/AIDS

Time-to-Death survival function defines survival times elapsed from treatment-initiation to death from AIDS regardless of treatment adherence. The Cox PH model was the default predictive model as it tested adequate with good-fit to the data to examine time-to-death survival times. The model’s global test alpha-level from Schoenfeld and scaled Schoenfeld residuals was highly nonsignificant [*p=*0.3168] and the cumulative hazard rates on the Cox-Snell’s curve run closer to the reference line implying good-fit to the data (Figure 3A) – in comparison to the Weibull model (Figure 3B); the best of the four parametric models.

**Figure 3:**
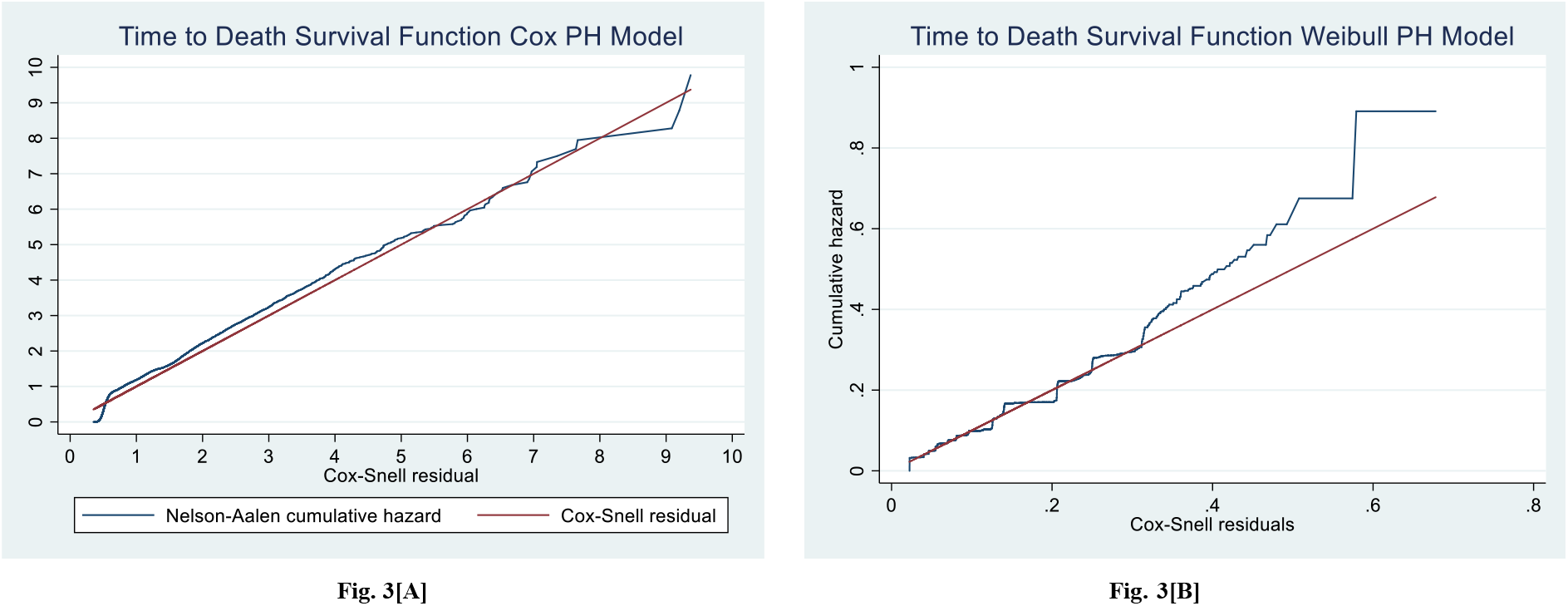
Goodness-of-Fit of Time-to-Death Survival Function Models. The Nelson-Aalen cumulative hazard rates run closer to the reference line on the Cox PH Model (**Fig. A**) than on the Weibull PH Model. Besides, the shapes of the Snell’s curves imply that the Cox PH is best-fitting the data.

In a univariate Cox PH regression analysis, crude hazard ratio findings demonstrated that demographic predictors: patients age and gender; geographical predictors: health facility’s locations; bioclinical predictors: ART eligibility, CD4+ cell count, Viral-RNA load, ART regimen type; aetiological factors, patients’ clinical presentation, HIV duration, ART period, were associated with time-to-death and risks (hazards) of death from AIDS.

In the multivariate Cox PH model, the analysis demonstrated the male gender was associated 42% increased hazard of death [aHR=1.42, (CI: 1.22–1.66)]. On patients age, increase in age of every 5 and every 10 years explained a 10% and 21% increased risk – such that, the hazard of death was increased by 63% in patients aged 20-39 years [aHR=1.63 (CI:1.09– 2.45], by 71% in patients aged 40-54 years [aHR=1.71 (CI: 1.14–2.57)] and by 2.7-fold higher risk in patients 55+ years [aHR=2.66 (CI: 1.09–3.53)] compared to those aged 15-19 years.

On geographical locations of health facilities, patients from Mzuzu Central and Thyolo District hospitals ART centres had 2.7-fold [aHR=2.66, CI:1.96–3.62) and 3.02-fold [aHR=3.02 (CI: 2.21–4.07)] higher risk of death respectively, compared to those from Chiradzulu ART facility. Moreover, subjects from semi-rural areas had 30% increased risk of death (*p<*0.001), however, the hazard was 20% inversely associated with urban areas [aHR=0.80, 95% CI: (0.69–0.91)], as was being single [aHR=0.86, 95% CI: (0.62–0.98)].

On patients symptoms at presentation to care, patients with chronic cough and/or breathlessness had 19% (aHR=1.19) increased risk of death, chronic diarrhoea or weight loss had 43% (aHR=1.43), chronic fever and/or severe headache had 30% (aHR=1.30), skin or oral lesion symptomatology had 33% (aHR=1.33) increased risks of death, compared to asymptomatic patients with low CD4 count and PMTCT’s Option-B+ presentations (*p<*0.004). These findings were reflected in aetiological factors analysis in which, in patients with ODs caused by infectious agents, the risks of deaths were significantly increased by margins of 31% to 76% (*p<*0.0001). Moreover, patients with WHO clinical stage III and IV at ART-initiation had 17.9-fold [aHR=17.90, 95% CI: 11.60–27.70)] and 20.1-fold [aHR=20.09 (95% CI: 13.30–32.70)] higher risks of death compared to asymptomatic: Option-B+ and CD4 count criteria patients.

On HIV surrogate markers: CD4+ count and viral-RNA load predictors, analysis findings demonstrated that with increase of every unit of baseline CD4 count; (1-cell/µL), the risk of death was reduced by approximately 1.0%. Equally, with increase in every 100 cells/µL of CD4 count, the risk of death reduced by 42% [aHR=0.58, (95% CI: 0.56–0.61)] and with increase of every 250 cells/µL the risk was reduced by 74% [aHR=0.26, (95% CI: 0.24–0.28). Hence, overall, the risk of death was increased by 17% in patients with CD4 count <250 cells/µL, compared to those with >250 cells/µL CD4 count [aHR=1.17, CI:1.08–1.23]. Subsequently, every 1-unit (1-c/mL) increase in baseline viral load (VL) was associated with approximately 1.0% increase in risk of death. Every increase in 1,000 copies/mL VL was associated with 0.0017%, and every increase of 10,000 copies/mL by 0.02% risk of death. The risk of death was therefore 6-fold higher, with every 1 million increases in VL [aHR=5.65, (CI: 5.25-6.08)]. Hence, patients with high baseline VL>1,000 copies/mL had 2.5-fold higher risk of death [aHR=2.46, 95% CI: (1.54–3.93)] compared to patients with baseline VL<1,000 copies/mL. Correspondingly, every increase of 100,000 copies, and 1 million copies of most recent VL levels were associated with 12% and 3-fold higher risk of death.

On ART regimes, the hazard of death was increased by 14% in patients commenced on Efavirenz-based first-line ART regimens compared to those commenced on Nevirapine-based therapies (aHR=1.14, CI: 1.00-1.30). Moreover, on HIV survivorship, the hazard of death was increased by 17%, 19% and 16% among patients with 3-5 years, 6-10 years and >10 years of living with HIV, respectively. On treatment initiation time variable, the findings demonstrated that with each passing year, the hazard of death was increasing by 8% [aHR=1.08, 95% CI: 1.04-1.12], compared to patients commenced on treatment in 2004 (*p*=0.013). In contrast, on HIV diagnosis year, with each passing year from 2005, the hazard of death decreased by 74% [aHR=0.26, 95% CI: 0.23-0.27], compared to patients diagnosed prior to 2004 (*p*=0.001).

## DISCUSSION

Survival times-to-death from HIV/AIDS has been extensively studied worldwide, but evidence from Malawi is lacking. The present analysis examined HIV survivorship from treatment-initiation to death in a non-concurrent, nationally representative retrospective cohort. Overall, death rates decreased with time and were statistically significantly associated with multiple factors as observed in this study.

This duration analysis of time-to-death survival function demonstrates that follow-up until December 2015 accounted for 24,105,125 person-days-at-risk (PDAR); females: 14,579,111 PDAR, males: 9,526,014 PDAR, and resulted in 11.8% (N=1,177) deaths; (females: N=686, males: N=491) from total sample (n=9,953) patients. Under this TTD survival function analysis, a total of 8,776 individuals were censored (did not experience the event), representing median survival time of 2,338 days (IQR: 1237 – 3353 days). The overall mortality rate was 1.78/100 person-years-at-risk (PYAR), with rates for females (1.72/100 PYAR) slightly lower than for males (1.88/100 PYAR). A total of 213 deaths (18.1%) occurred early in year-one post-ART-initiation. Most deaths occurred in young adults of 20-39 years (N=470, 39.97%), and mid-aged adults of 40-54 years (N=483, 41.07%), and were mostly due to mycobacterial conditions (N=106, 37.3%) in particular TB infection (N=103) most of which were PTB cases (N=69, 66.9%). The proportion of death was high in Southern region (63.13%, N=743) while Northern region had least deaths (N=313) (*p<*0.0001).

Based on Kaplan Meier curves, survival of patients decreased gradually from the beginning then tailed-off at about 7,500^th^ survival days to reach its minimum value (62.5% survival) at the end of the follow-up (9,224^th^ survival day) [Figure 4(a)]. Overall, survival was slightly higher in females especially from about 3,900^th^ survival day, to nearly the entire data generation period (except in final months) [Figure 4(b)]. The estimated survival function however, do not reach zero, implying that the greatest observed survival time in this study was a censored value [23].

**Figure 4:**
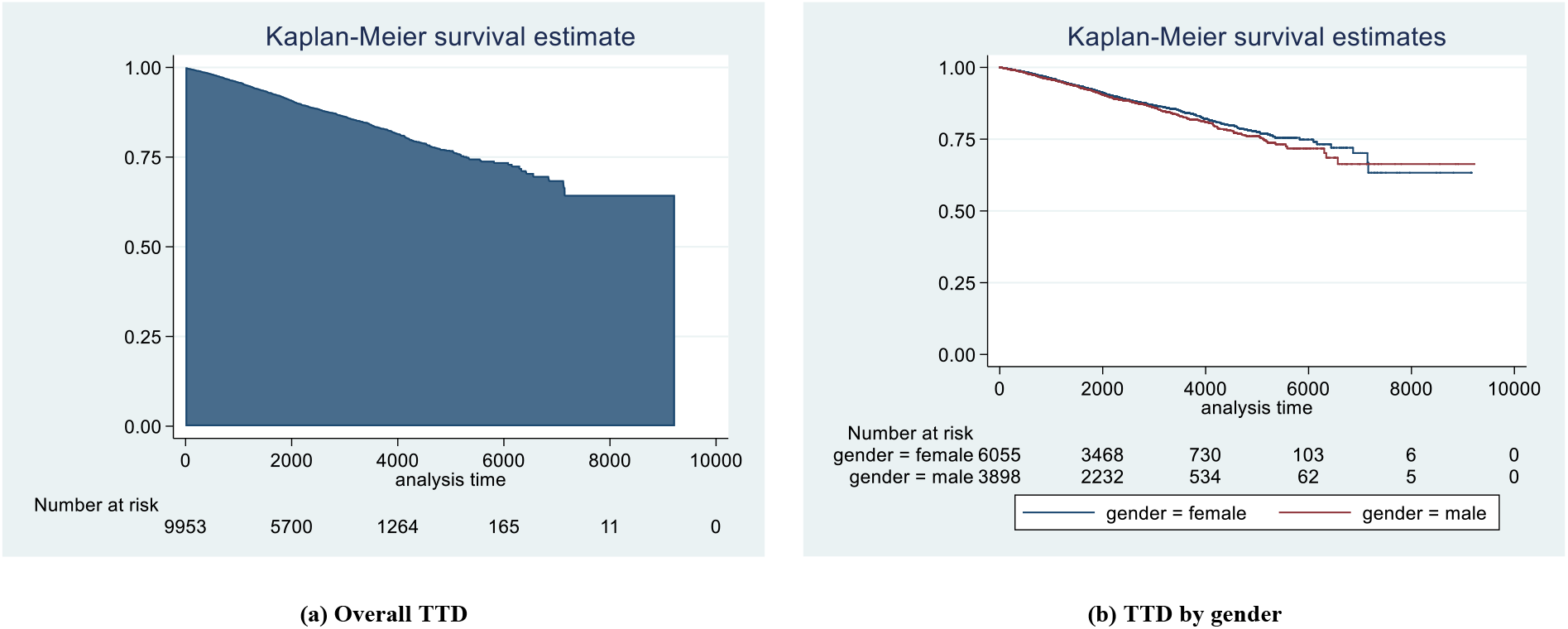
Kaplan-Meier Survival Function Curves of Overall Time-to-Death and by Gender. Figure 4[A] shows that overall, the survival of patients gradually decreased from the beginning then tail-off at about 7500^th^ survival days to reach minimum value (of 62.5% survival) at the end of follow-up (9224^th^ survival day). In figure 4(b), the survival estimates have been stratified by gender and the curve demonstrates that survival was slightly higher in females especially from about 3900^th^ survival day to nearly the entire 11-year data generation period except the final months. Then estimated survival do not reach zero – implying a sensored value.

HIV mortality during year-one follow-up post-ART-initiation was 18.1% (N=213). This rate was lower than from Ethiopia studies which reported year-one death rates between 56% and 62% [4, 24-26]. Besides, higher death rates during first months of ART were also reported from prospective cohorts in South Africa and Tanzania [27, 28]. In a multivariate Cox PH model adjusted for ART eligibility, demographical, socioeconomical, geographical, and bio-clinical factors were significantly associated with time-to-death survival function.

### Demographic Predictors Associated with Time-To-Death from HIV/AIDS

The multivariate findings demonstrated that male patients had 42% higher rate of death than females. This gender difference could be related to active health seeking behaviours among females which enable them to know their HIV serostatus early and initiate treatment timely relative to males; a factor also demonstrated from other studies. The significance of gender in relation survival times until death has been widely studied and produced similar findings to the present study. A retrospective study from Tanzania reported a 4.7-fold higher risk of death in males; [aHR=4.71] compared to females [29]. Similar findings were also reported from China [30], and Ethiopia [31-33]. Moreover, a UK study inversely reported 27% reduced risk of death [aHR=0.73] among female HIV patients compared to male counterparts [34].

The present study also demonstrated that age was linked to increased hazard of death. The findings show that with every 1-year increase in age, the risk of dying was correspondingly increasing by 1%. And increase in age of every 5- and 10-years explained 10% and 21% increased risk of death, respectively. Upon factoring the ‘age’ variable, hazard of death was observed increasing across all age groups with rates ranging from 61% to as high as 3-fold (aHR=2.66) especially in older patients. The age bracket in this study constitutes the economically productive Malawi population. Hence, increasing mortality in this group has had adverse implications on national productivity and developments [35, 36]. The present study’s findings are consistent with others across the globe which demonstrated that high age bracket has higher risk of death and that the risk increases with ageing. Studies from Ethiopia reported a 4-fold and 10-fold higher risk among 35+ year-olds [4, 33], a 3-fold-increase in 35-44 year-olds [37], and >2.5-fold higher risk among 45+ year-olds [38]. Furthermore, from Tanzania, a 4% increase was reported with any increase in 1-year age [39], while from Nigeria, 82.0% increase in mortality was reported among 21-50-year-olds [40], and from China a 4-fold higher risk among 50+ year-olds was reported [30]. The UK study describing mortality and their causes in PLHIV in the HAART-era reported huge increases in hazard of death associated with advancing age especially among 45+ years-olds [34]. Thus, patients aged 65+ years had 11-fold higher hazard risk of death; [aHR=11.33]; similar but higher result than 2.6-fold from Spain [41], 4.5-fold from Hong Kong [42] and 2.4-fold from Zimbabwe [43]. In this present study, being single was the only predictor statistically significantly associated with reduced hazard of death: [aHR=0.86] – a rather an inconsistent finding with most studies including one from Ethiopia which demonstrated no association [5].

### Socioeconomic and Geographic Predictors Associated with Time-To-Death

Under socioeconomic status, self-employment was associated with 12% reduced risk of death from HIV. Likewise, secondary education was associated with 15% reduced risk of death. Comparative evidence for these predictors is however lacking from literature.

On geographical associations with risk of death, compared to rural areas, semi-urban residence was associated with 30% increased hazard of death (aHR=1.30%) whereas urban residence was associated with 20% reduced risk (aHR=0.80). hence, Thyolo District Hospital; a semi-urban, secondary level ART centre in Southern region of Malawi, had a 3.02-fold higher risk, and Mzuzu Central hospital; an urban, 3^rd^ level ART centre in Northern region, had a 2.66-fold higher risk of death, respectively, compared to Chiradzulu hospital; another semi-urban, secondary tier, health facility in Southern region. The findings explain (to some extent), existing challenges facing Malawi health system such as chronic shortage of resources including health professionals and medical supplies among others [44]. The geographical status findings in this study appear consistent with those from Northwest Ethiopia which reported 2-fold higher hazard of death (aHR=2.18) in patients attending ART services in hospitals as opposed to those in health centres [38]. However, the findings were inversely related to those from Zimbabwe [43].

### Bioclinical Predictors Associated with Time-To-Death

The bioclinical predictors analysis showed that all five infectious aetiological agents responsible for ODs were associated with increased hazard of death by 31% to 76% (aHR=1.31 to 1.76). However, the non-infectious neoplastic factors demonstrated no association. This finding strengthens the proposition that while HIV-seropositivity is the sole strong precursor for death, coinfection with any infectious pathogenic agent more than doubles and highly increases one’s risk of death. The same is true with neoplastic conditions comorbidity despite the present study data falling short of the finding. In addition, it also demonstrates that infectious opportunistic diseases, other than neoplasms, are responsible for most HIV-related death. To my knowledge, association of aetiological factors with time-to-HIV mortality has not been documented before; hence, comparisons are drawn from evidence on specific conditions (from similar aetiological agents) which have been widely studied.

To this end, the findings demonstrated that some patients’ clinical features present at first presentation to care were highly associated with increased hazard risk of death. Thus, breathlessness and/or coughing, chronic diarrhoea and/or weight loss, chronic fever and/or headaches, skin and/or oral lesions, were highly associated with increased risk of death from AIDS– reflecting and supporting the findings in aetiological agents’ analysis. The study findings on the association of clinical features with time-to-death are consistent with findings from Tanzania which found that chronic diarrhoea, chronic cough, fever, skin lesions, dysphagia, and oral lesions, were significantly associated with increased risk of death among HIV/TB coinfection adults [39].

The hazard risk of death was also increased among patients with AHD. Thus, WHO clinical stage III & IV of HIV disease explained a 17.8-fold [aHR=17.90] and 20.09-fold [aHR=20.09] higher hazard risk of death, respectively, reflecting the immunosuppressive nature of HIV infection. This finding is consistent with a large body of global scientific evidence demonstrating that AHD is a strong precursor for death in PLHIV [4, 5, 33, 37-43, 45, 46].

Similarly, the role of HIV surrogate markers; CD4 count and viral load, in association to death has been widely studied. In the current study, low CD4 count, and high VL, were associated with increased risk of death in that CD4 count <250 cells/µL was associated with 17% increased rate of death whereas, unsuppressed VL >1,000 copies/mL was associated with 49% increased risk of dying from HIV. The study findings; especially on CD4 count, are consistent with studies from other regions including Ethiopia [4, 5, 47], Vietnam [46], Hong Kong [42], Korea [45], Spain [41], India [48] and Zimbabwe – with an inverse finding whereby high CD4 count (>50 cells/µL), was associated with reduced hazard of death [43].

The present study also examined the impact (efficacy) of different antiretroviral drug (ARV) classes on HIV-related deaths (on preventing AIDS mortality). The ARVs are regrouped Nevirapine-based, Efavirenz-based, boosted-Ritonavir-Protease inhibitors and non-standard (NS-ART) regimes. The findings demonstrated that the hazard risk of death increased by 14% if Efavirenz-based regimens were used as first-line therapies. The finding, however, is incongruent to the Korean retrospective study which determined mortality causes and risks of death in HIV patients but failed to find significant association between ART classes; PI-based, NNRTI-based, or Mixed regimens, and death [45]. This predictor however appears not to have been widely studied as evidence is scanty. Our findings could be explained by the fact that although ARVs have been and continue to be successfully used in treating HIV, their action remains to prolong life other than to cure the disease. Besides, ARVs are (often) toxic (and are not without adverse effects) – especially earlier lines upon which substantial proportion of this study subjects were commenced on.

On ART period predictor, findings showed that late-ART period (2011-2015), was associated with 4-fold increased hazard of death (aHR=3.53). This finding was clinically unexpected as treatment during this period was commenced in relatively less immunosuppressed patients with CD4 cell count <500 cells/µL who should have had better survival chances. Arguably, it has been established that a proportion of individuals who start treatment with CD4 counts <350 cells/µL do not achieve CD4 counts >500 cells/µL up to after 10 years on ART [49, 50]. Besides, their life expectancy is shorter than those who initiate ART at higher CD4 count thresholds [49-51]. This, therefore, spells out the importance of following recommended guidelines including active follow-up of new treatment-naïve patients.

As with other illnesses, HIV seropositivity substantially increases one’s risk of death especially due to its progressive immunosuppression especially when untreated. HIV duration variable analysis was aimed to examine potential hazard of death along HIV survivorship period, i.e., against the progressive period of living with HIV. Findings from this analysis demonstrated that 3-5 years, 6-10 years, and 10+ years of HIV survivorship, was statistically significantly associated with 27%, 39%, and 56% higher risk of AIDS death compared to <2 years of HIV duration. However, the predictor likewise appear exceptional to the ADROC study as previous evidence is missing.

ART-initiation-year variable explored annual proportions of patients newly initiated on treatment (annual-ART-naïve patients). The findings demonstrated that risk of death progressively increased with each additional year throughout the 2004 to 2015 data generation period. Thus, patients who initiated treatment in 2004 and 2005 had low but increasing hazard of death: (aHR=1.32 & 1.76 respectively), whereas those initiated in 2011 & 2012 had a 6.6-fold and 9.28-fold higher risk of death: (aHR = 6.60 & 9.28 each). However, patients commenced on treatment in 2014 and 2015 had substantially higher risk of death; 15-fold (aHR=15.43 & 15.04 each). A similar analysis was reported in Korean study, but the findings are incongruent as they observed no association [45]. On the other hand, no significant results were observed in annual new HIV cases predictor despite significant findings in univariate model. This finding is similar to Spanish study which also reported no significant findings [41]. The reason for progressive increase in hazard of death under this analysis could not be explained however, could be correlated to improved reporting.

## CONCLUSION

The study has shown demonstrated factors that likely to affect and impact HIV survivorship and contribute to the higher HIV mortality in Malawi. Overall, the study revealed high mortality in this population especially among the productive persons age -groups. The overall median estimated survival time of this sub-cohort was 2,338 days (i.e., 6.4 months). The study has shown high mortality of the cohort in the first year post-ART-initiation of up to (N=213 deaths) especially in earlier months of treatment. The result of Cox PH predictive model showed that both host and nonhost factors can be significant predictors to survival times in of HIV/AIDS patients. The goodness-of-fit of baseline distribution using graphical method and Cox-Snell residuals plots revealed that Cox distribution is better model than the parametric Weibull distribution to explain survival times HIV/AIDS patients data.

## Data Availability

All data produced in the present study are available upon reasonable request to the corresponding author.

## Authors’ contributions

HHS contributed 100% of research work, thus; Conceptualization, Data curation, Methodology, Formal analysis, Writing – original draft, Writing – review & editing, Writing – final draft.

## Acknowledgements

The author is grateful to all gate keepers and research teams from all 8 research sites: Nkhoma Hospital, Lighthouse Trust & Martin Preus Centre, Partners-in-Hope, Mzuzu Hospital, DREAM Centre, Chiradzulu Hospital, QECH, and Thyolo Hospital ART facilities for their undivided support towards this study.

## Availability of data and materials

All data for the study are available from the corresponding author on reasonable request.

## Ethical approval and Informed consent to participate

Study’s ethical approval was granted by the University of Warwick Research Ethics Committee; The Biomedical and Scientific Research Ethics Committee (BSREC), and from Malawi Research Ethics Committee; The National Health Sciences Research Committee (NHSRC) in the Ministry of Health. The need for consent to participate was waived-off due to nature and source of data.

## Supplementary materials

None.

